# Population-based analysis of knee joint loading in a knee osteoarthritis cohort: the impact of PCA-derived gait kinematic variations on estimated medial knee contact forces

**DOI:** 10.1101/2024.01.31.24301806

**Authors:** Giacomo Di Raimondo, Miel Willems, Bryce Adrian Killen, Sara Havashinezhadian, Katia Turcot, Benedicte Vanwanseele, Ilse Jonkers

**Affiliations:** Department of Movement Sciences, Katholieke Universiteit Leuven, 3001 Heverlee, Belgium; Department of Kinesiology, Université Laval, Québec, QC G1V 0A6, Canada

## Abstract

Osteoarthritis (OA) is a prevalent musculoskeletal condition leading to functional limitations, especially among the elderly. Current treatments focus on pain relief and functional improvement, however there is a lack of approaches which slow disease progression. A promising approach focusses on reducing knee joint loading, as excessive loading contributes to knee OA progression. This study explores kinematic variations in the knee OA population, utilizing principal component analysis (PCA) to examine gait variations (primitives) in both healthy individuals and those with knee osteoarthritis (KOA) and their implications for knee joint loading. The KOA population exhibited 14 modes of variation representing 95% of the cumulative variance, compared to 20 in the healthy population, indicating lower variability with KOA. The relation between identified gait primitives and knee loading parameters, revealed complex relationships. Surprisingly, modes with the largest kinematic variations did not consistently correspond to the highest variations in knee loading parameters revealing degrees of freedom which may have a larger role in determining joint loading. Moreover, potential gait-retraining strategies for KOA, associating specific kinematic combinations with altered knee loading were identified. The results showed a good agreement with previously applied strategies. However, this study highlights the importance of analyzing whole-body kinematics for effective gait retraining, as opposed to focusing on one single joint variation. The study’s insights contribute to understanding the intricate interplay between gait pattern variations and knee joint loading changes in healthy and KOA populations, offering practical applications for guiding interventions and estimating loading parameters.

## 1. Introduction

Osteoarthritis (OA), is a prevalent and complex musculoskeletal condition, accounting for a large societal and economical burden on both individuals and healthcare systems worldwide. It is considered a major cause of impaired functionality, especially among the aging population [1]–[3] and is associated with joint pain and stiffness, leading to reduced mobility. While age is a risk factor, other mechanical factors such as obesity, joint misalignment, and overuse also play crucial roles in disease development and progression. With an aging population and lifestyle changes, the prevalence of knee OA is increasing, posing substantial socioeconomic challenges [2], [3]. At present, conservative knee OA treatments, focusing on improving knee function and slowing down joint damage via pharmacological and/or physical therapy approaches are missing [4]. As such, effective interventions to slow down disease progression represent an unmet clinical need, that, if successful, offer evident societal and economic benefits [2]. Avoiding mechanical risk factors by e.g. reducing knee joint loading has been suggested to be a promising strategy to slow down disease progression [5]–[7]. Indeed, physiological knee joint loading is needed to preserve structural integrity for absorbing and distributing loads during motion and plays a crucial role in maintaining cartilage homeostasis. Alterations to either the magnitude and/or location of knee joint loading have been suggested to contribute to degeneration [8], [9]. Moreover, factors like trauma, misalignment, or joint instability are recognized as contributing factors that can induce changes in knee movement. These changes can consequently lead to further alterations in knee joint loading associated with osteoarthritis [10], [11].

Gait kinematics in healthy and knee OA populations have extensively been studied [12]–[16]. Specifically, patients with KOA reported decreased walking speed and step length, reduced range of motion (ROM) in knee flexion (6–10°) but increased ROM in knee adduction (2-8°) compared to healthy individuals [17], [18]. Furthermore, aberrant joint moments have been reported whereby the knee adduction moment (KAM) and ankle adduction moment were (up to 19%) higher compared to healthy individuals. Additionally, hip, knee and ankle flexion moments were reported to be reduced in the knee OA group [17]. These previously reported alterations in gait pattern have been suggested to likely induce altered knee joint loading [13], [19]–[21] in this population.

Direct measurement of knee joint loading *in-vivo* requires invasive measurement techniques via instrumented knee implants that measure the forces transmitted through implants [22]–[24]. However, this approach is costly, invasive, and not feasible in large cohorts. Thus, musculoskeletal (MSK) model-based simulations based on 3D motion capture (MoCap) data represent the state-of-the-art method to estimate *in-vivo* joint loading [24], [25]. Several groups have reported good agreement between MSK model-based and instrumented knee implant joint loading parameters [26], [27], hence making it a feasible alternative to instrumented prosthesis [26], [28] with the advantage of being able to be applied to larger clinicial cohorts.

Using these MSK modeling-based workflows, literature specifically reports differences in loading distribution in the medial and lateral knee compartments in the KOA population. Knee joint loading is frequently increased compared to the healthy population (up to 22%), with elevated loading rates [29] and an imbalanced load distribution that results in increased loading of the medial compartment. This, in turn, leads to excessive cartilage loading and potential damage [27], [28]. Moreover, an association between elevated knee contact forces during walking and the three-year radiographic progression of knee osteoarthritis has been demonstrated, which underlines the importance of mitigating knee load by reducing the compressive knee contact forces [30].

While the above-mentioned studies are useful in documenting differences in kinematics and in knee loading distribution between the two populations (healthy and KOA), they are based on small sample size and cross-sectional data. Consequently, the sampled knee joint loading landscape does not account for the overall variability in the gait pattern. Furthermore, no causal relation between gait variability and knee joint loading can be established. This is indeed relevant as gait pattern variability constitutes coordinated movements of different segments of the legs and trunk, which each by themselves but also in combination will affect knee joint loading. The gait pattern adaptations, which likely also constitute compensatory mechanisms for avoiding knee pain for instance, may in fact worsen knee joint loading and hence disease state.

Therefore, it is essential to improve the current cross-sectional data on knee joint loading by incorporating insights from natural variations in gait patterns observed in both healthy individuals and those with KOA. This can be achieved by integrating population-based analysis of gait kinematics with MSK modeling to provide a more comprehensive understanding of the landscape of knee joint loading. Principal component analysis (PCA) has been previously used to enhance motion characterization at a population scale, identifying primary gait pattern variations [31]–[34]. Whereas already characterized in healthy individuals, such analysis in patients with knee OA is currently non-existent, leaving it open if primary gait variations exist in knee OA. Furthermore, in combination with MSK modeling, PCA would allow to investigate how dominant variations in gait pattern affect knee joint loading. Investigating the gait patterns variations within populations and their impact on knee joint loading can potentially define movement primitives (i.e., the main joint kinematic variations observed in the population) that predispose to altered knee joint. As gait retraining strategies are a feasible and increasingly popular therapeutic approach in patients with KOA, this analysis may offer the conceptual basis for defining the gait characteristics to target in order to impact knee joint loading and hence impact pain and functional impairments [35], [36].

This study presents a population-based analysis of joint kinematics and knee joint loading using PCA-based gait primitives of healthy and knee OA individuals to (1) identify primary kinematics variations in healthy and knee OA populations and determine if these variations are unique to each population and (2) investigate how these primary gait pattern variations contribute to changes in knee joint loading parameters. Our hypothesis is that given the observed cross-sectional disparities between knee OA and healthy gait patterns, primary modes (principal components) of variation would differ between these two populations and that we can identify crucial movement characteristics able of reducing knee loading in a knee OA. These unique movement characteristics can serve as the foundation for gait retraining to slow or stop the progression of knee OA.

## 2. Results

### 2.1 Gait pattern variations

We identified the gait primitives underlying the gait characteristics of KOA and healthy individuals. To this end, the modes (principal components) that represent 95% of the observed cumulative variance in each population were considered for further analysis. Twenty modes were required for the healthy population, but only 14 modes for knee OA population, potentially indicative of reduced gait variability in the knee OA population (Fig. 1 and Tab. S1). Based on the selected modes, we reconstructed kinematics during an entire gait cycles for each of the considered 14 degrees of freedom (i.e., a gait pattern representative of each mode). This allows us to identify specific kinematic variations compared to the mean gait pattern, which are representative of the specific mode. Figures 2 and 3 show examples of two different reconstructed gait patterns from a single mode, in this case, mode 1.

**Figure 1:**
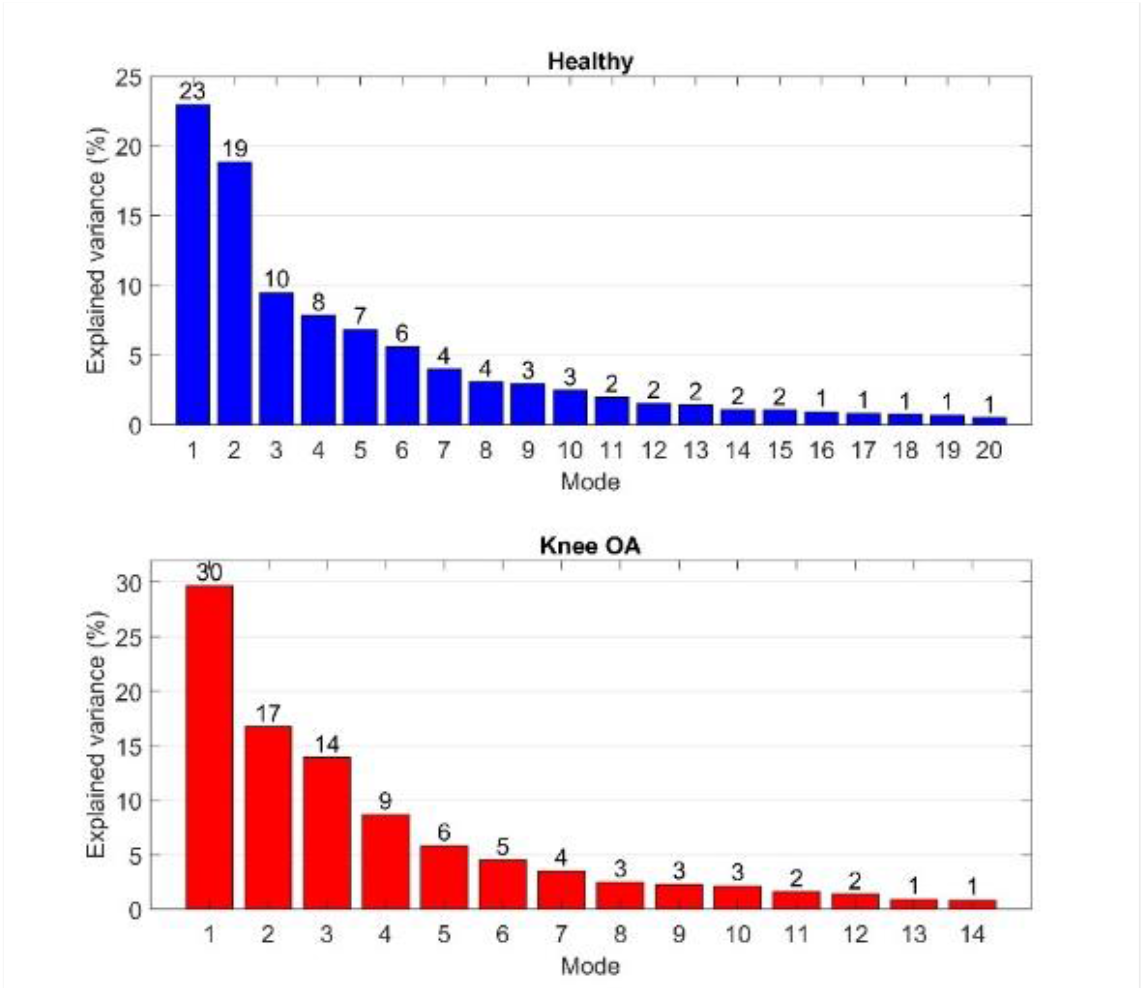
Explained variance (%) of individual modes that represented a 95% cumulative explained variance of the population variation obtained from Principal Component Analysis (PCA) of 2553 gait cycles for healthy (top - blue) and 1756 gait cycles for KOA (bottom-red)

**Figure 2:**
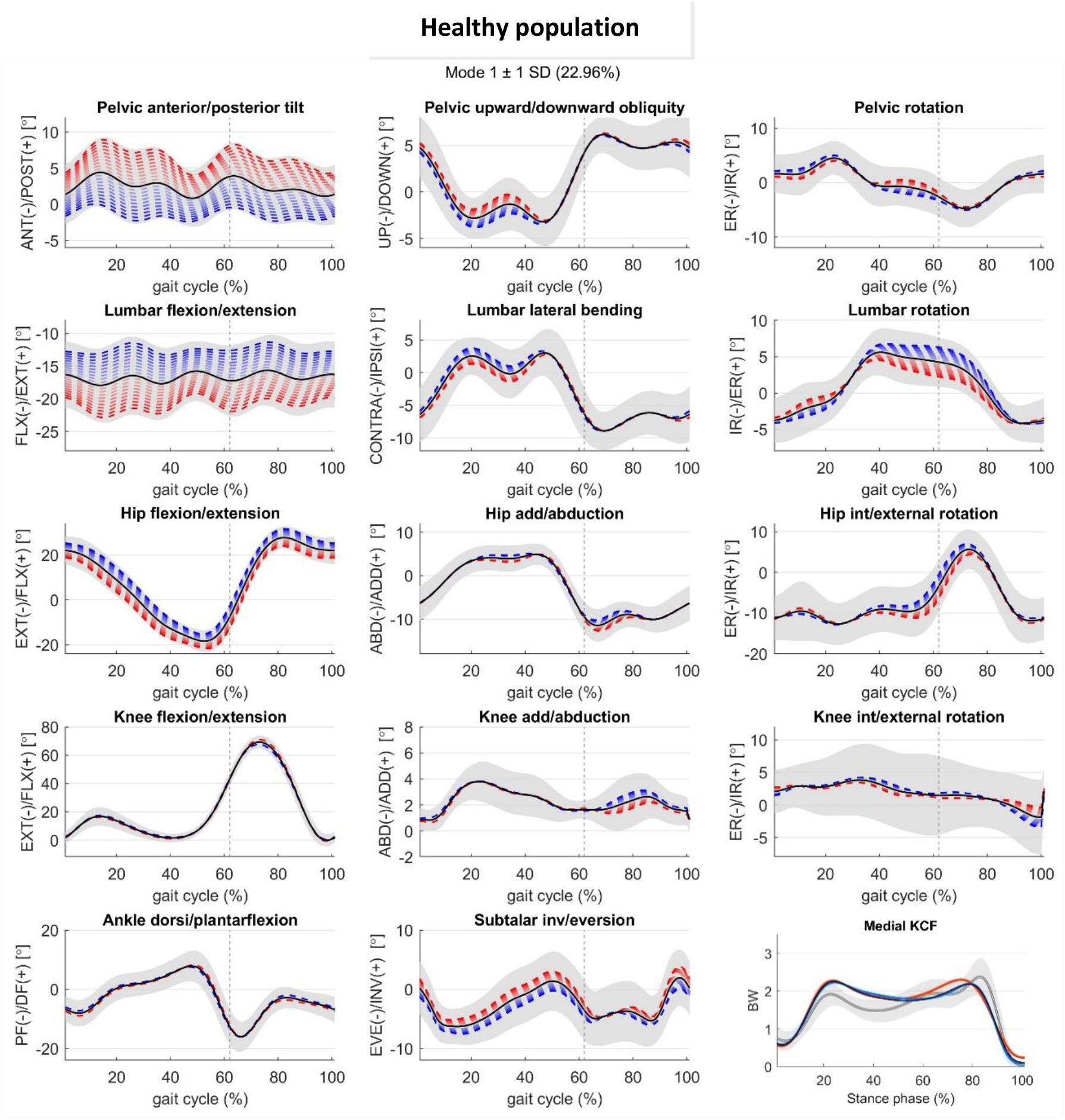
Joint kinematic variations of the reconstructed gait pattern described by Mode 1 (22.96% of explained variance) with +/-1 standard deviation from the mean gait pattern with positive (red), and negative (blue) standard deviation for healthy population dataset – ipsilateral leg. Solid black line shows the mean of the dataset and the grey regions shows the spread of the input dataset (2553 gait cycles), and grey dashed line defines the stance phase. Medial knee contact force (KCF) (bottom right panel) assessed based on mean (black) and Mode 1 +/-1 standard deviation (red, blue respectively) kinematic patterns. Mean and standard deviation of measured knee contact forces in healthy OA population (grey solid line and area).

**Figure 3:**
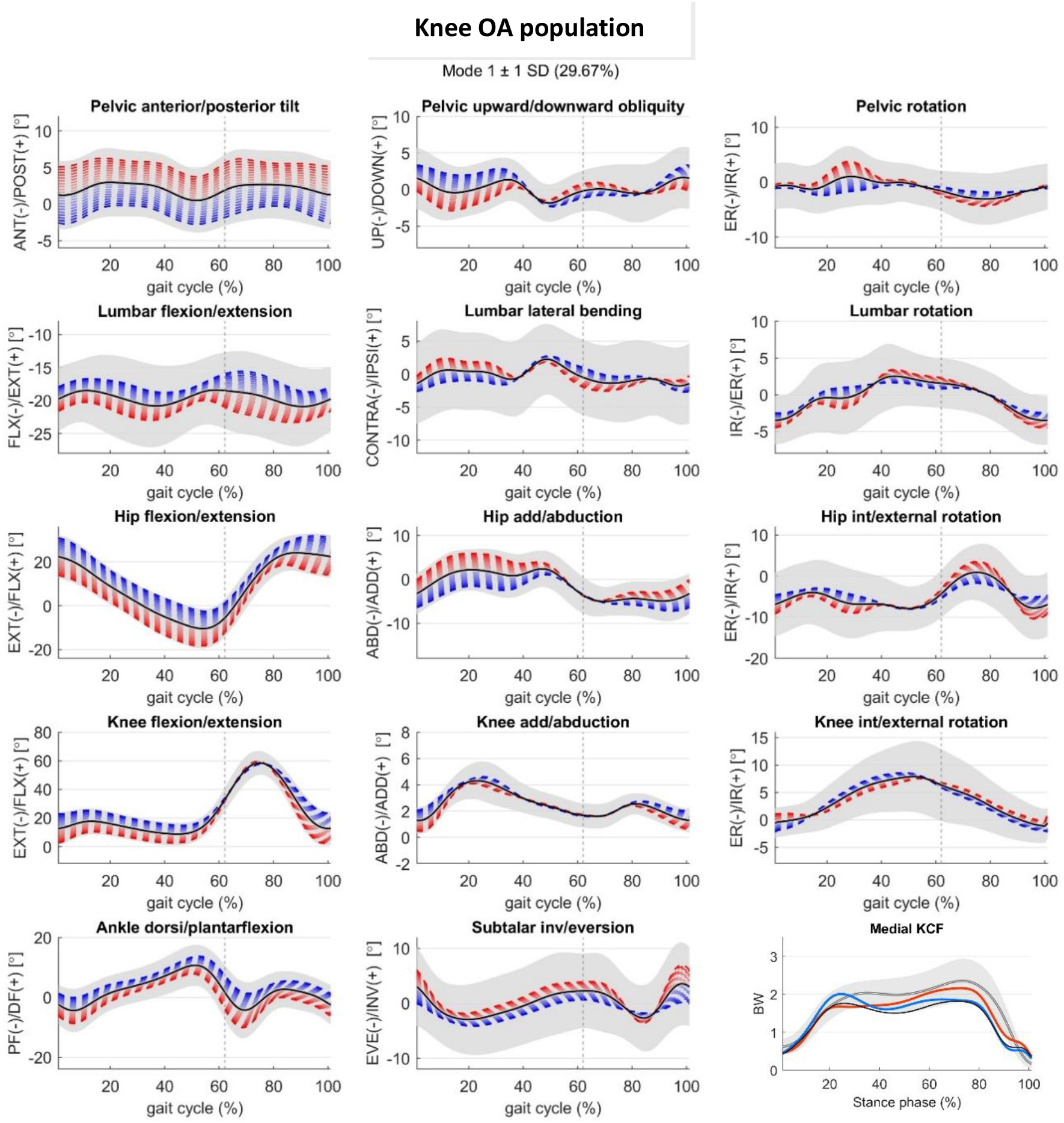
Joint kinematic variations of the reconstructed gait pattern described by Mode 1 (29.67% of explained variance) with +/-1 standard deviation from the mean gait pattern with positive (red), and negative (blue) standard deviation for knee OA population dataset – affected leg. Solid black line shows the mean of the kinematic dataset, the grey regions show the spread of the input dataset (1754 gait cycles), and grey dashed line defines the stance phase. Medial knee contact force (KCF) (bottom right panel) assessed based on mean (black) and Mode 1 +/-1 standard deviation (red, blue respectively) kinematic patterns. Mean and standard deviation of measured knee contact forces in knee OA population (grey solid line and area).

We subsequently summarized the contribution of each individual degree of freedom (DOF) to each mode’s variation (Figure 4) by illustrating which DOFs are responsible for the first 50% of entire mode variation. Mode 1, accounting for 23% and 30% of the total variation in healthy and in knee OA individuals respectively, is associated with changes in sagittal plane angles in both populations, however there are distinct joints contribution. Specifically, it reveals that in both populations, the highest variation is observed in hip flexion/extension and pelvic anterior-posterior tilt movements. The third joint differs between the populations; it involves lumbar flexion/extension for the healthy population but knee flexion/extension for the knee OA population. Mode 2, accounting 19% and 17% of the total variation in healthy and in knee OA individuals respectively, is primarily associated with hip int/external rotation and knee flexion/extension in both populations. However, the relevant joint DOFs start to differ, introducing hip flexion/extension and knee int/external rotation for healthy and lumbar flex/extension, lumbar lateral bending and subtalar joint for knee OA. Mode 3, accounting for 10% and 14% of the total variation in healthy and knee OA individuals respectively, introduces ankle dorsiflexion/plantarflexion in both populations. In the healthy population, knee internal/external rotation becomes more relevant, while in the knee OA population, the subtalar joint. Further analysis of the different joints identified as most involved for each subsequent mode (4 to 20 – less than 8% and 9% of the total variation in healthy and knee OA individuals respectively, see Fig.1,) revealed that overall that the kinematic variations are quite distinct with a different number of joints.

**Figure 4:**
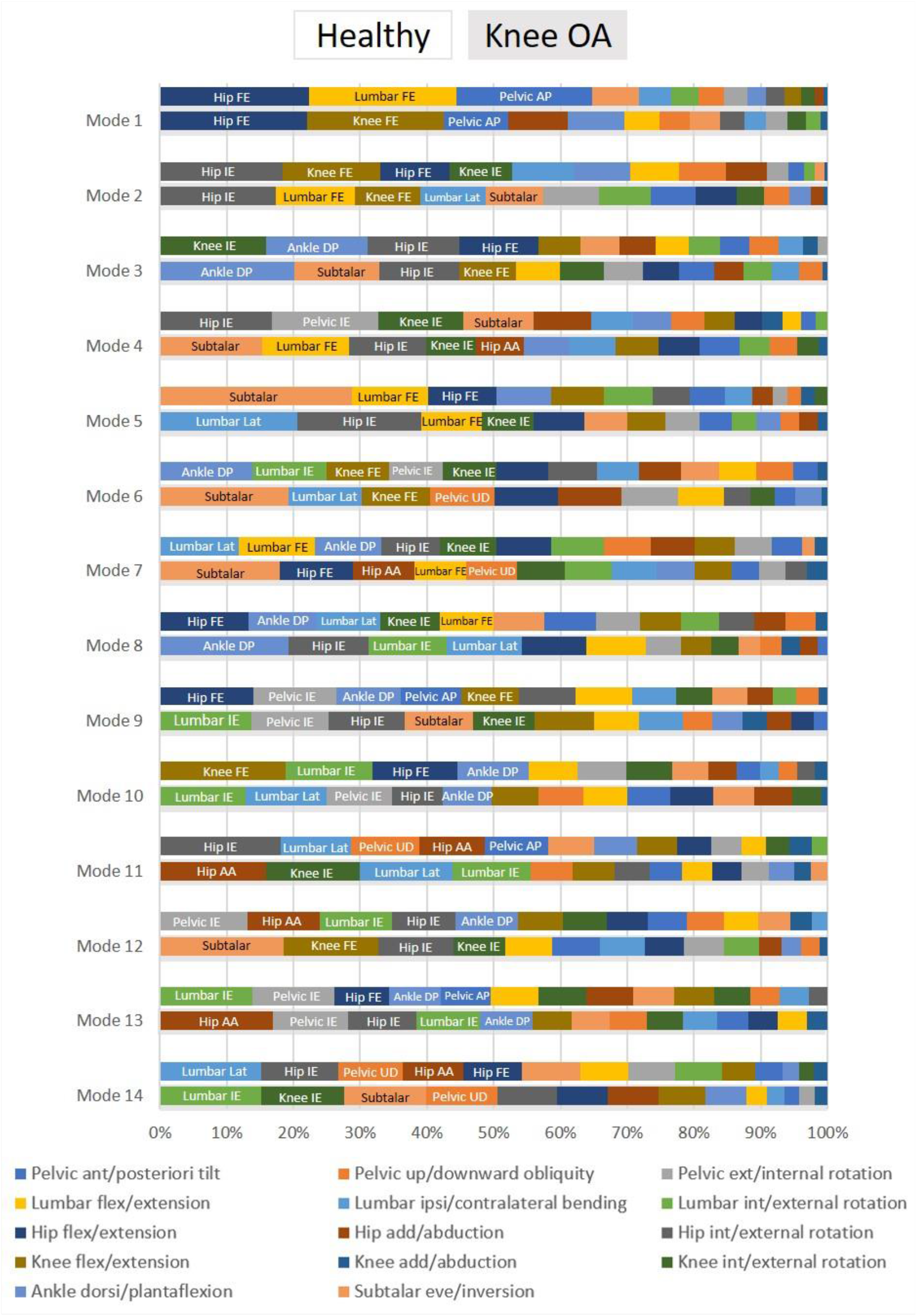
Comparison of joint variations contribution percentage (1-100%) of each mode over stance phase for healthy (white background) and knee OA (grey background) populations. Each bar color represents a different joint degree of freedom and its width denotes its contribution (%) to the kinematic variation of the mode. Only joints that contribute to the top 50% (when sorted from largest to smallest) are highlighted with text. Mode 15 to 20 for healthy population are illustrated in supplementary materials (Fig. S1).

### 2.2 Knee contact force variations

We investigated the effect of the observed gait primitives on estimated knee contact force defined by the modes from the healthy and KOA population. Specifically, each single unique gait pattern for each unique mode was used as input in a previously developed MSK modeling workflow [37] to estimate the knee contact forces (Fig 2 and 3 bottom right panel). As such, we can examine how the joint variations illustrated in Fig. 4 affect the estimated contact force peaks in the two populations (Fig. 5 and 6) with the intention of defining relevant changes in knee contact force peaks (increases/decreases). For simplicity, we described only the modes that resulted in functionally relevant changes in contact forces peaks (changes exceeding ±10% BW, see Methods section). Modes with functionally relevant medial contact force peak changes are illustrated in Figures 5 and 6. Complete modes with medial and lateral contact forces peaks changes are illustrated in supplementary materials (Fig. S2, S3).

**Figure 5:**
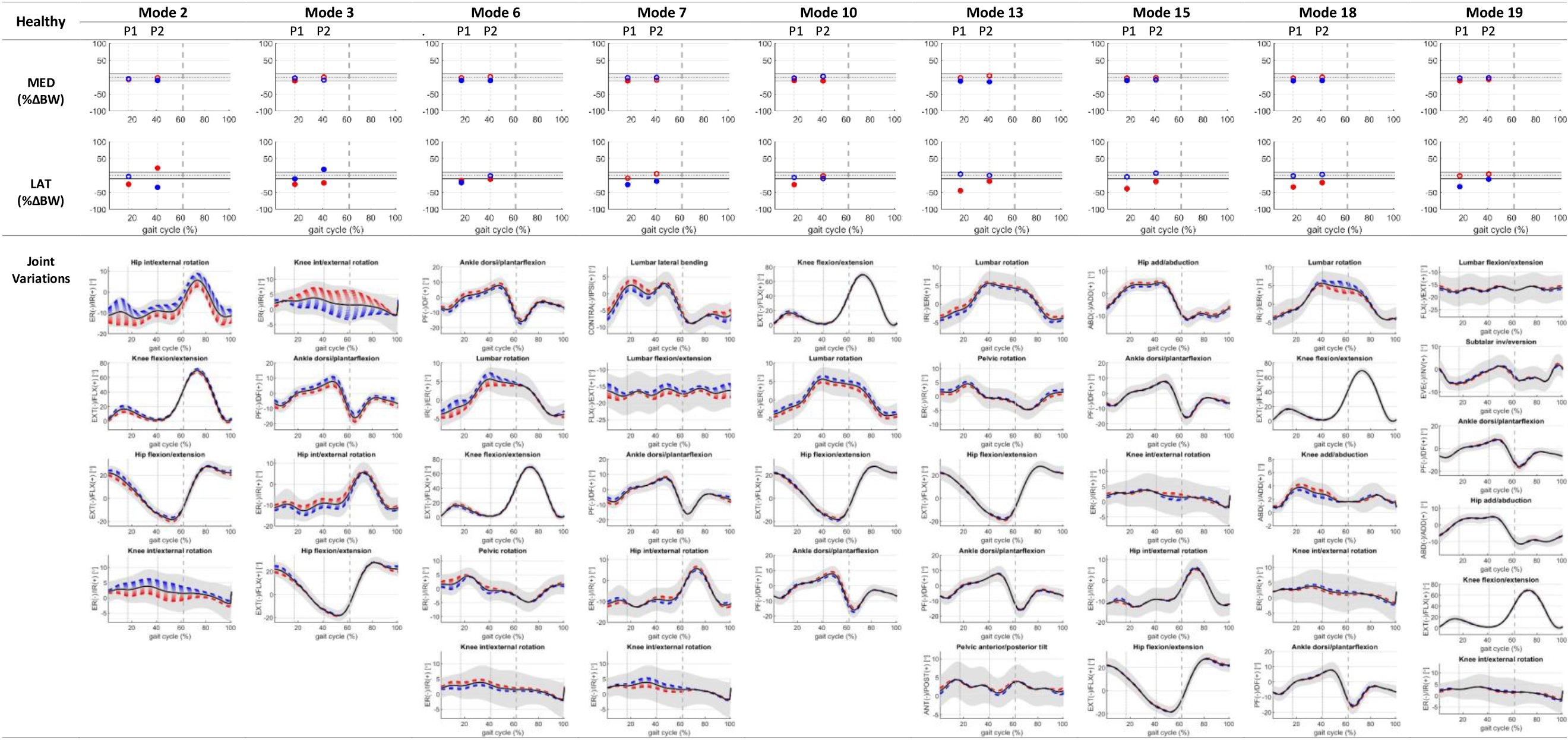
Functionally relevant changes as result of kinematic variations in medial compartment knee contact forces and relative changes in lateral compartment (relevant changes are depicted with filled red and blue circle) expressed as a % difference in body weight (BW) compared to knee contact forces estimated using the mean healthy gait pattern (black dotted lines), at the top. Solid grey lines are the cut-off/threshold for functionally relevant knee contact force changes (see Material and Methods section 4). Below (bottom) are the kinematics of the different joints contributing to the top 50% (see Fig 4) of the observed variation for each specific mode depicted as ±1 STD (red, blue respectively). Grey dashed line defines the stance phase.

**Figure 6:**
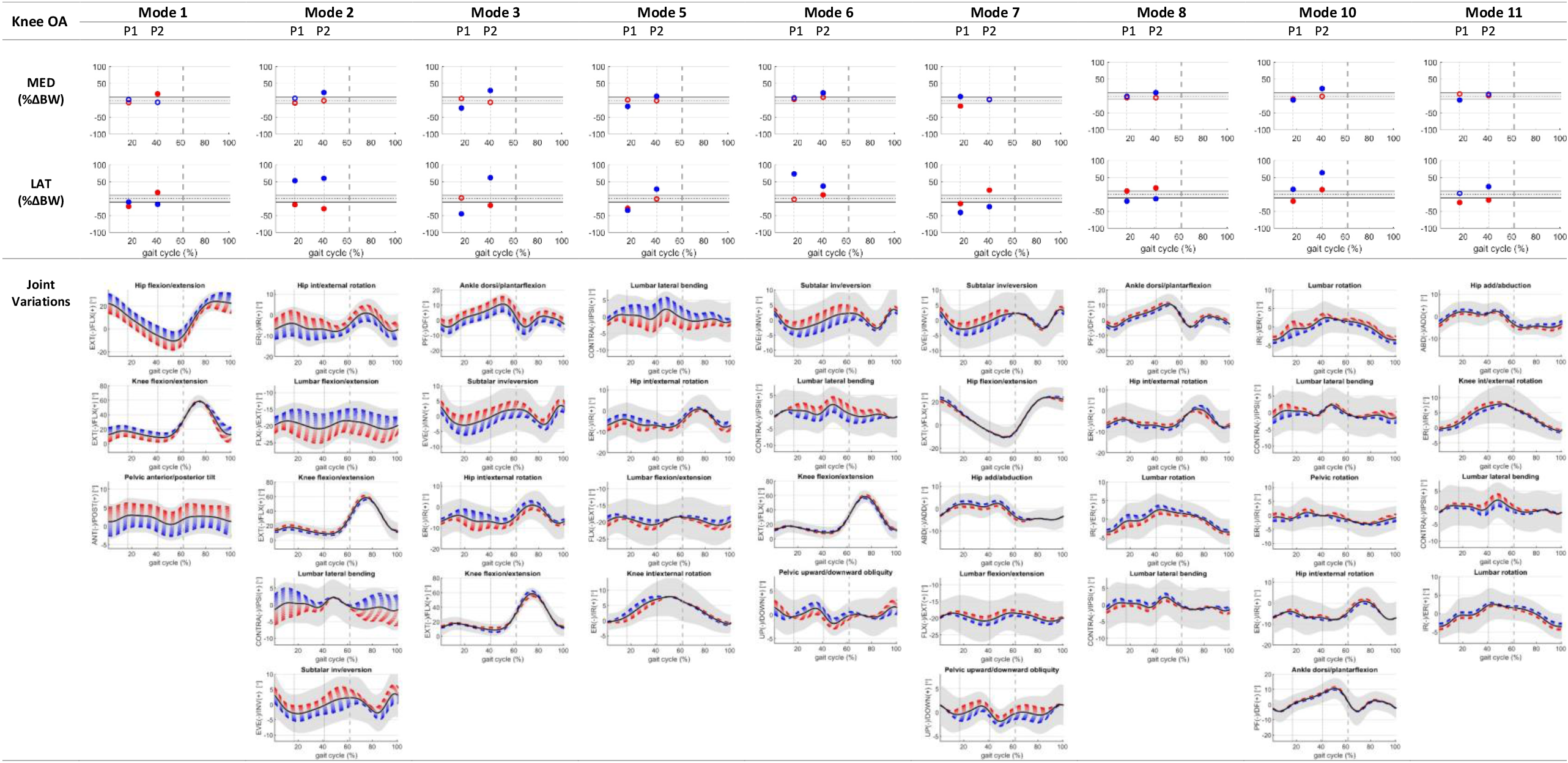
Functionally relevant changes as result of kinematic variations in medial compartment knee contact forces and relative changes in lateral compartment (relevant changes are depicted with filled red and blue circle) expressed as a % difference in body weight (BW) compared to knee contact forces estimated using the mean knee OA gait pattern (black dotted lines), at the top. Solid grey lines are the cut-off/threshold for functionally relevant knee contact force changes (see Material and Methods section 4). Below (bottom) are the kinematics of the different joints contributing to the top 50% (see Fig 4) of the observed variation for each specific mode depicted as ±1 STD (red, blue respectively). Grey dashed line defines the stance phase.

#### 2.2.1 Healthy population

##### Gait primitives/kinematics affecting medial compartment loading

Overall, no functionally relevant **increases** (>10% BW) in contact force were observed for either the 1st or 2nd peaks in any of the modes. However, a number of different modes showed a functionally relevant **decrease** (<10% BW) in either or both the first and second peaks of knee joint loading. In the analysis, joints were categorized based on the number of associated modes. The ankle joint, specifically ankle plantarflexion (modes 3, 6, 7, 10, 13, 15, 18, 19), exhibited the most common involvement across healthy population modes. The hip joint, including hip flexion (modes 2, 13, 15), hip extension (modes 3, 10), hip internal rotation (modes 2, 3, 15), was observed in multiple modes, along with hip external rotation (mode 7), hip abduction (mode 15), and adduction (mode 19). This suggests that the influence the hip joint on medial contact forces may be modulated by the combination with other joints. This is evident from the observation that alterations in any degree of freedom (DOF) of the hip, irrespective of direction (i.e., flexion or extension, internal or external), can lead to a decrease. The knee joint, including knee extension (modes 6, 10, 18), and knee external rotation (modes 6, 7, 15, 19), also showed functionally relevant variations across different modes. The lumbar joint, including lumbar internal rotation (modes 10, 13), lumbar external rotation (modes 6, 18), along with lumbar extension (mode 19) and ipsilateral bending (mode 7), showed functionally relevant variations. The pelvis degrees of freedom, specifically pelvic internal rotation (mode 13), external rotation (modes 6), and anterior tilt (mode 13), were present but less frequently across the relevant modes. Subtalar eversion was observed only in mode 19.

#### 2.2.2 Knee OA population

##### Gait primitives/kinematics affecting medial compartment loading

In the knee OA population, different modes demonstrated functionally relevant **increases** (>10% BW) in either or both the first and second peaks of knee contact forces. Notably, the hip joint exhibited the most common involvement across various modes, including hip extension (modes 1, 3, 7), internal rotation (modes 3, 5, 8), external rotation (mode 2, 10), flexion (modes 7), abduction (mode 7), and adduction (mode 7). The lumbar joint showed consistent involvement across various modes, including lumbar ipsilateral bending (modes 5, 8, 10), contralateral bending (mode 2, 6), extension (mode 2, 5), external rotation (8), and flexion (7). This implies that, like in the healthy population, the impact of the hip and, in this case, the lumbar joint as well, on medial contact forces is affected by the combination with other joints, irrespective of the movement direction. The knee joint, including extension (modes 1, 2, 3), internal rotation (mode 5), and flexion (6), also exhibited functionally relevant increases in knee joint loading across different modes. Similar to the healthy case, pelvic joint movements such as internal rotation (mode 10), downward obliquity (6), upward obliquity (mode 7), and posterior tilt (mode 2) were present but less frequently across the relevant modes. In contrast to the healthy population, where the ankle joint showed the most common involvement across various modes, the knee OA population exhibited relevant increases primarily during ankle plantarflexion (mode 3, 8, 10). Furthermore, subtalar eversion was observed in mode 3, 6 and 7.

Functionally relevant **decreases** (<10% BW) in either or both the first and second peaks of medial knee contact forces were observed. The hip joint exhibited the highest involvement across various modes, including hip extension (modes 3, 7), internal rotation (modes 3, 5), external rotation (mode 10), and abduction (mode 7); however, hip flexion and adduction did not show any decrease in knee joint loading. The lumbar joint, including lumbar ipsi/contralateral bending (modes 5, 10) and flex/extension (modes 5 and 7), showed notable decrease in knee joint loading. Similarly to the increase case, hip and lumbar joints contributed to reducing loading when combined with other joints depending on their direction. The knee joint showed functionally relevant decreases across different modes, including knee extension (modes 1, 2, 3), and flexion (mode 6). Notably, in the case of knee internal rotation (mode 5), a similar reduction in loading was observed, highlighting a consistent trend across various modes. Examining the pelvic joint, combination of internal rotation (mode 10) and downward obliquity (mode 7) showed always relevant decreases in knee joint loading. The ankle joint demonstrated less involvement across various modes compared to the healthy population, including only plantarflexion in only two modes 3 and, 10). Additionally, subtalar eversion was observed in mode 3 and 7, respectively. This discrepancy in joint involvement across modes underscores how gait pattern variations impact loading changes are different in the two populations, emphasizing the distinct ways in which joints contribute to altered loading patterns.

### 2.3 Joint combinations associated to relevant loading changes

The results showed relevant increases or decreases in contact force peaks in both healthy and knee OA populations, based on variations in kinematic patterns defined by the assessed modes (Fig. 5 and 6). However, upon focusing on functionally relevant changes in medial contact force peak alterations irrespective of population, interestingly, some recurrent kinematic pattern variations emerge, inducing consistently an increase or decrease in medial knee contact force peaks (Fig. 7).

**Figure 7:**
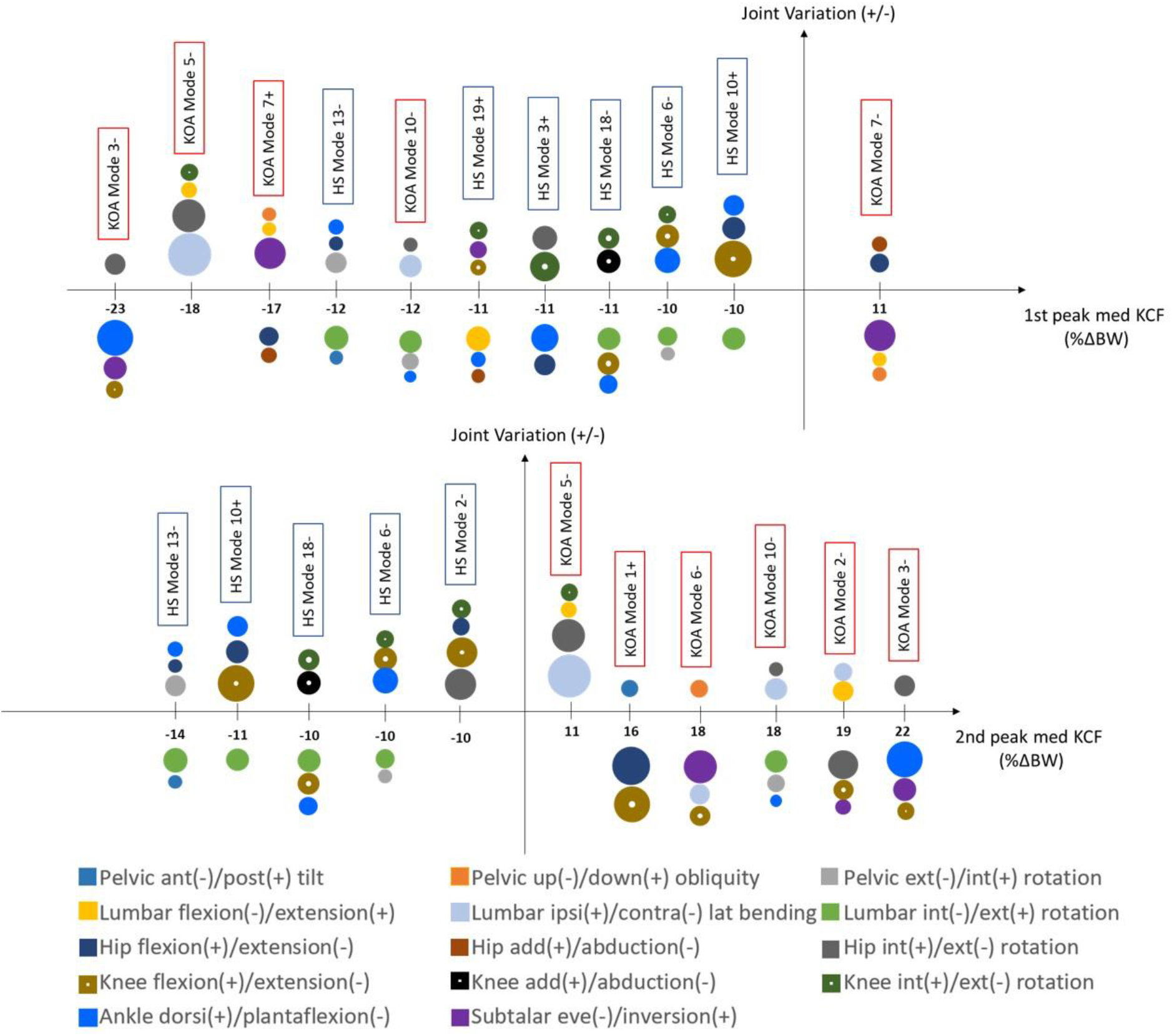
Functionally relevant changes as result of kinematic variations in first (top) and second peak (bottom) medial compartment knee contact forces (KCF) expressed as a % difference in body weight (BW) compared to knee contact forces estimated using the mean gait pattern. Each colored bubble represents a different joint degree of freedom and its dimension denotes its contribution (%) to the kinematic variation of the mode with ±1 STD (HS: healthy in blue label; KOA: knee OA in red label). Only joints that contribute to the top 50% are shown as illustrated in Fig. 4.

It was observed that combinations of joint variations with lumbar internal rotation (light green) consistently leads to a decrease in both medial peaks, and combinations with lumbar external rotation lead to an increase on the second peak. Combinations with knee flexion (light brown) consistently resulted in peak decreases; however knee extension, which mainly leads to second peak increases, could lead to decrease if combined with lumbar internal rotation (light green) and/or ankle plantarflexion (bright blue). Combinations of knee internal rotation (dark green) consistently resulted in decreases in both peaks. In addition, subtalar eversion and inversion (purple) consistently leads to increases and decreases in both peaks, respectively. However, other kinematic variations, such as hip int/external rotation (dark grey) and lumbar ipsi/contralateral bending (light blue) (i.e. found to have a functionally relevant impact on medial knee loading changes in modes 2 and 3), influence changes in contact force peaks depending on the combination with other joints. These combinations can either increase or decrease medial contact forces peaks.

## 3 Discussion

This study aimed to identify population-specific kinematic variations or so-called gait primitives in healthy or knee OA populations and how they affected knee joint loading. To this end we applied PCA to a large set of gait kinematics data from both healthy and knee OA population combined with a previously developed MSK modelling workflow [37].

Unique gait primitives i.e gait kinematics variations were identified for each population. Knee OA population showed 14 modes of variation compared to the 20 of the healthy population, showing higher variability. Mode 1, accounting for the highest total variability (23% for healthy and 30% for knee OA), was associated with variations in hip flexion/extension and pelvic anterior/posterior tilt for both populations. In addition, lumbar flexion/extension was observed in healthy, while knee flexion/extension in knee OA, highlighting differences between the two populations. Notably, despite the knee OA population, showing reduced range of motion (ROM) in the knee kinematics (approximately 6–10 degrees) compared to healthy population, (Fig, 2 and 3), similar to previous studies [12], [17], [21], [38], it showed higher knee flexion/extension variability in mode 1 (Fig. 3). This suggests a larger relevance for the knee OA population than the healthy one in the primary mode. An interesting observation is that, as we advance through the modes, mode 2, 3, 4 etc., an increasing number of degrees of freedom (DOFs) are required to account for a cumulative 50% variation for each mode (Fig. 4), varying distinctly between the two populations. This suggests that a greater combination of degrees of freedom (DOFs), involving up to 6 joints, is necessary to characterize each mode, where potentially the changes in all DOFs are closer together, rather than having one or two DOFs being distinctly dominant (i.e. mode 1, 2).

The second objective of this study was to investigate how the joint variations defined by the different modes in the two populations contributed to changes in knee joint loading parameters (i.e. contact forces peaks). Interestingly, modes presenting the largest variations in kinematics (i.e. hip flexion/extension, pelvic tilt in mode 1) did not translate into modes presenting the highest variation in knee joint loading parameters. In fact, mode 2 and mode 6 in knee OA with total explained variance of 17% and 5%, respectively, showed the highest increase for both medial contact force peaks (6% for peak 1 and 23% for peak 2). Mode 7, with total explained variance of 5%, showed the highest decrease for medial contact forces (17% for peak 1 and 3% for peak 2). Surprisingly, Mode 3, with total explained variance of 14%, revealed a maximal decrease in peak 1 (23%) but a maximal increase in peak 2 (29%) of medial contact forces. Moreover, we observed that these later modes require more degrees of freedom (DOFs) to account for 50% (Fig. 3), and it appears that relatively smaller changes in joint DOFs (via PCA) have a larger impact compared to a single or a few DOFs, as seen in earlier modes of variation (i.e. mode 1, 2). This highlights once again that considering variations in the whole-body kinematics when examining loading changes, instead of focusing solely on a single joint variation, is crucial.

Interestingly, opposite changes in kinematics (i.e. knee flexion or extension) did not consistently result in opposite changes in knee joint loading parameters. For instance, both combinations of knee flexion and ankle dorsiflexion (mode 6) and knee extension and ankle plantarflexion (mode 18) corresponded to relevant decreases in knee loading in healthy population. However, joint variation combinations with lumbar internal rotation or knee flexion or knee internal rotation resulted in relevant decreases, and combinations with knee extension or subtalar eversion consistently resulted in relevant increases in contact forces peak (i.e. mode 6 10, 18, Fig. 7). However, other joint combinations with hip internal/external rotation and lumbar ipsi/contralateral bending resulted in either increased or decreased medial contact forces peaks dependent on the specific kinematics combination (i.e. mode 2, 5, Fig. 7).

Based on the population-based analyses, specific movement characteristics could be identified that are already targeted in currently implemented gait-retraining strategies [13], [39]. For example, previous studies have demonstrated that trunk leaning, particularly ipsilateral trunk leaning, can decrease medial knee contact force peaks [20], [40]–[44]. Indeed, we observed relevant changes in contact force peaks associated with modes related to lumbar flexion/extension or ipsi/contralateral bending (i.e., mode 4 and 5 in knee OA), suggesting their potential utility in gait retraining. While these variations were confirmed in modes 5 and 6, it’s important to note that in some cases, they could result in either increased or decreased medial contact force peaks dependent on the combination of whole-body kinematics (i.e mode 5, Fig 7). Also other modes associated with combinations in ankle dorsi/plantarflexion, subtalar inversion/eversion and hip internal/external rotation (i.e. mode 3, Fig. 7) showed relevant contact force peak increases/decreases. This described joint combination can be associated with changes in foot progression angle, toe-in, toe-out strategies. These alterations have been previously associated with significant changes in knee moments (flexion and adduction moment) and contact force peaks [21], [45], [46]. Overall, our findings align with literature, providing consistency with reported gait pattern modification outcomes. This strengthens the potential practical application of our study’s findings in guiding and validating specific gait retraining interventions. However, additional relevant changes were observed in mode 15 for healthy population and in mode 11 for knee OA associated with variation in hip adduction/abduction during walking, which did impact the distribution of knee loading between the medial and lateral compartments. It has been shown that walking with increased hip abduction, which is moving the foot outward, can lead to higher knee loading on the medial compartment [44]. Potentially, based on our results, combinations involving either lumbar internal rotation (mode 10) or knee internal rotation (mode 5) and knee flexion (mode 3) may be further explored as strategies to decrease knee contact force peaks.

The population-based kinematics variations could potentially serve as a foundation for defining the combined joint variations for healthy and knee OA populations that lead to either reduced or excessive knee loading conditions [20], [47]. Importantly, this study results showed that knee loading changes are dependent on the whole-body kinematic variations. Consequently, it is crucial not to only consider gait pattern modifications in isolation at the single-joint level. Gait retraining should consider changes in multiple joints. Indeed, the described modes do not exclusively define changes in one degree of freedom; instead, they define combined kinematic changes in all joints, involving variations in pelvic, lumbar, hip, knee, and ankle joints [48]. Therefore, evaluating the impact of whole-body kinematics on knee joint loading with MSK modeling and expanding biomechanics measurement systems for use in more ecological contexts is crucial to evaluate the effect execution of the whole body kinematic strategy needed to effectively reduce knee loading.

In our study, we limited the description of each mode to a restricted number of degrees of freedom (up to 6), making it is a highly selective approach. A more comprehensive understanding of what each mode describes would require a detailed comparison of whole-body kinematics, which is a limitation of our study. Moreover, it was observed that the knee OA population exhibited a greater variation in knee contact forces, with both relevant increases and decreases across various modes (Fig. S4). This variation showed a range of up to 32%, in comparison to the healthy population where only decreases of up to 21% were observed. As such, our PCA analysis fails to identify specific kinematic patterns in the healthy population that could lead to functionally relevant increases in medial contact forces. Finally, gait data were collected during treadmill walking to facilitate the acquisition of a large number of trials. Treadmill walking kinematics are known to differ subtly from overground walking. This should be considered upon interpretation of the identified kinematic strategies and transferring them to overground conditions.

In conclusion, this study offers valuable insights into the relationship between gait pattern variations and knee joint loading changes in healthy and knee osteoarthritis populations. Through principal component analysis, specific movement characteristics associated with altered knee loading were identified, highlighting unique gait characteristics and loading changes for both populations. The findings demonstrate the importance of analyzing the whole gait kinematics to optimize knee loading reduction, taking population biomechanical factors into account. Moreover, the study suggests the potential use of reconstructed gait patterns (PCA-based) to estimate loading parameters. This innovative approach opens up possibilities for future applications, particularly through the utilization of machine learning techniques to accurately predict distinct gait patterns and establish correlations with knee loading variations during everyday activities. This not only promises a practical and easily accessible solution but also represents an alternative to complex data collection in motion capture laboratories and musculoskeletal modeling workflows.

## 4 Material and Methods

### Dataset

3D MoCap collected at Laval University, Quebec City, Canada from twenty-three healthy adults and seventeen patients diagnosed with medial knee OA walked on instrumented treadmill at self-selected speed for ∼2 min (between 65 and 167 gait cycles per participant – 2553 and 1754 cycles in total for healthy and knee OA population, respectively).

**Table 3:** Demographic of the dataset used. KL: Kellgren-Lawrence.

|  | Healthy | Knee OA |
| --- | --- | --- |
| **Gender** | 10M / 13F | 5M / 13F |
| **Age (yr)** | 36.5 ± 4.3 | 66.6 ± 7.3 |
| **Height (m)** | 1.72 ± 0.08 | 1.65 ± 0.12 |
| **Mass (kg)** | 73.3 ± 4.6 | 80.5 ± 15.3 |
| **BMI (kg/m<sup>2</sup>)** | 24.62 ± 4.55 | 29.86 ± 6.28 |
| **KL grade** | 0 | 2-4 |
| **Speed (m/s)** | 1.28 ± 0.13 | 0.75 ± 0.23 |

The 3D position of the 74 reflective markers, 42 attached to anatomical landmarks of the different body segments [49], [50] and 32 cluster markers was recorded using 9 infrared camera system (VICON, Oxford Metrics Group, UK, 100Hz) while walking on an instrumented treadmill (Bertec, Columbus, OH, US, 1000Hz) recording ground reaction forces. All participants provided written informed consent, prior to data collection. This research was in accordance with the ethical guidelines provided by the ethical research committee Centre Intégré Univeritaire de santé et de services sociaux de la Capitale-National, Quebec (MP-13-2020-1954).

### Data processing

Filtered, labelled and gap-filled MoCap data (marker trajectories, ground reaction forces and center of pressure) were exported as .trc and .mot files from Nexus 2.12. Joint kinematics were calculated using the inverse kinematics tool in OpenSim Joint Articular Mechanics (JAM) using a validated musculoskeletal model with combined 12 degrees of freedom for the tibiofemoral (6DOF) and patellofemoral (6DOF) joints [28]. All trials were processed and time-normalized to 100% of the gait cycle (101 time points) using custom-built MATLAB scripts. Moreover, OpenSim JAM [51], [52] with the integrated concurrent optimization of muscle activations and kinematics (COMAK) algorithm [52] was used to solve the muscle activation distribution problem, compute the resultant secondary knee kinematics and knee contact forces [26] for all measured data.

### Data analysis

After completing the data processing, joint kinematics were parameterized using a PCA-based framework [19], [34], [53]. Principal Component Analysis (PCA) of 2553 gait cycles for healthy and 1756 gait cycles for KOA were separately applied to extract the mean as well as dominant features in the modelling kinematic curves. Only the modes that represented > = 95% of the population variation were considered (20 for healthy and 14 for knee OA) for further analysis and MSK modelling (Table S1). Gait patterns were reconstructed using the different modes with +/-1 standard deviation from the mean gait pattern. (i.e. Fig. 2 and 3). This approach allowed for a targeted and isolated analysis of each mode and their contribution to the knee loading parameters. Each reconstructed gait pattern represented by each individual mode +/-1 standard deviation were utilized as input in a previously developed pipeline that integrated probabilistic principal component analysis (PPCA) [37], [54] and the zero moment point (ZMP) methods [55], [56] to estimate ground reaction forces, moments (GRFM), and the center of pressure (COP) and subsequently estimate the knee joint loading parameters (pressure, contact force, contact area) [26], [28], [52]. Thus, the impact of the reconstructed gait patterns, represented by each individual mode, on the change in knee contact force (KCF) peaks was investigated. Relevant changes in knee contact force peaks were reported if greater than ±10% body weight difference (ΔBW) from the mean knee contact force peaks of the medial knee compartments. In general, previously reported differences in KCF peaks between healthy and KOA population were found to be at least 10% BW [57], [58].

## Author Contributions

Conceptualization, G.D.R., B.A.K., B.V., I.J.; Data collection, S.H. and K.T.; Data curation, M.W. and G.D.R.; Formal analysis, G.D.R, B.A.K.; Funding acquisition, K.T. and I.J.; Investigation, M.W., G.D.R. and B.A.K.; Methodology, G.D.R., B.A.K.; Software, G.D.R.; Supervision, B.A.K., B.V. and I.J.; Validation, G.D.R., and B.A.K.; Writing original draft, G.D.R., B.A.K. and I.J.; Writing—review and editing, M.W., B.A.K., B.V. and I.J. DeepL (https://www.deepl.com/write) was used in the writing process to improve readability and language writing. All authors have read and agreed to the published version of the manuscript.

## Institutional Review Board Statement

The ethical committee of Centre intégré universitaire de santé et de services sociaux de la Capitale-Nationale, Québec (MP-13-2020-1954) gave ethical approval for this work.

## Informed Consent Statement

Informed consent was obtained from all individuals involved in the study.

## Data Availability Statement

The workflow and data will be made available on RDR—the KU Leuven repository.

