## Supplementary materials for "Population-based analysis of knee joint loading in a knee osteoarthritis cohort: the impact of PCA-derived gait kinematic variations on estimated medial knee contact forces"

***Table S1****: Modes that represented up to 95% cumulative variance of the population variation obtained from Principal Component Analysis (PCA) of 2553 gait cycles for healthy and 1756 gait cycles for KOA.*

| Mode N | Healthy (%) | | Knee OA (%) | |
|---|---|---|---|---|
| | Explained variance | Cumulative variance | Explained variance | Cumulative variance |
| 1 | 22.96 | 22.96 | 29.67 | 29.67 |
| 2 | 18.84 | 41.80 | 16.72 | 46.39 |
| 3 | 9.49 | 51.29 | 13.91 | 60.30 |
| 4 | 7.84 | 59.13 | 8.68 | 68.98 |
| 5 | 6.83 | 65.96 | 5.81 | 74.79 |
| 6 | 5.61 | 71.57 | 4.51 | 79.30 |
| 7 | 3.99 | 75.55 | 3.52 | 82.82 |
| 8 | 3.09 | 78.64 | 2.48 | 85.30 |
| 9 | 2.94 | 81.58 | 2.27 | 87.57 |
| 10 | 2.46 | 84.04 | 2.14 | 89.70 |
| 11 | 1.96 | 86.00 | 1.59 | 91.30 |
| 12 | 1.52 | 87.52 | 1.38 | 92.68 |
| 13 | 1.43 | 88.95 | 0.92 | 93.60 |
| 14 | 1.08 | 90.04 | 0.82 | 94.42 |
| 15 | 1.05 | 91.09 | - | - |
| 16 | 0.93 | 92.02 | - | - |
| 17 | 0.83 | 92.85 | - | - |
| 18 | 0.74 | 93.60 | - | - |
| 19 | 0.69 | 94.29 | - | - |
| 20 | 0.50 | 94.79 | - | - |

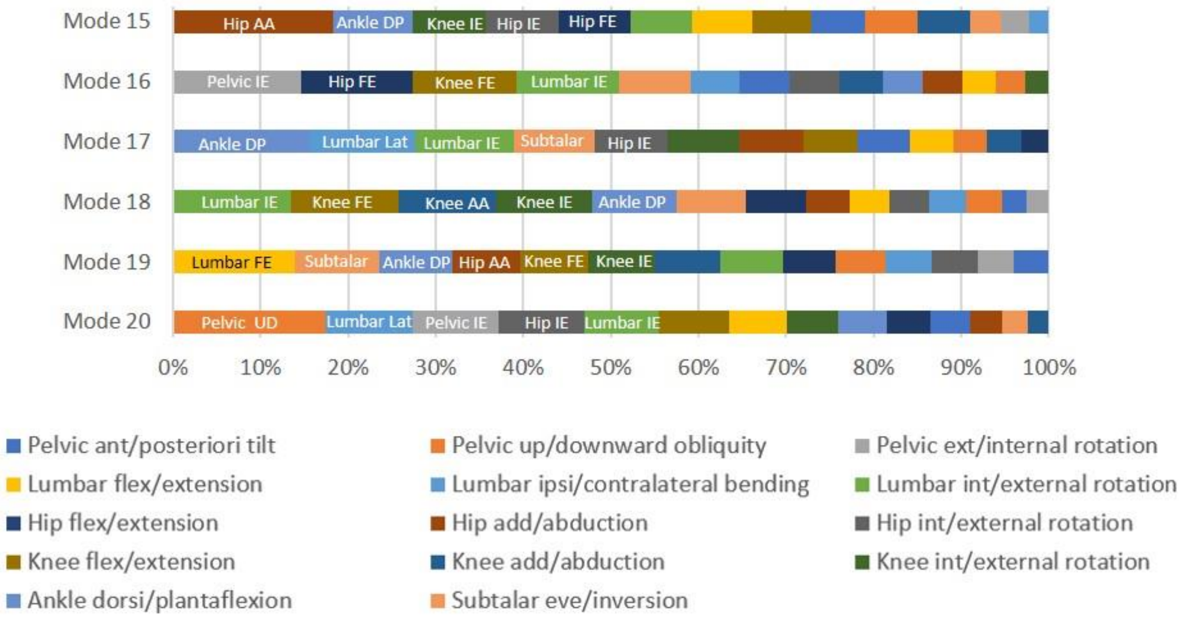


***Figure S1****: Comparison of joint variations contribution percentage (1-100%) of mode 15 to 20 over stance phase for healthy population. Each bar color represents a different joint degree of freedom and its width denotes its contribution (%) to the kinematic variation of the mode. Only joints that contribute to the top 50% (when sorted from largest to smallest) are highlighted with text.*

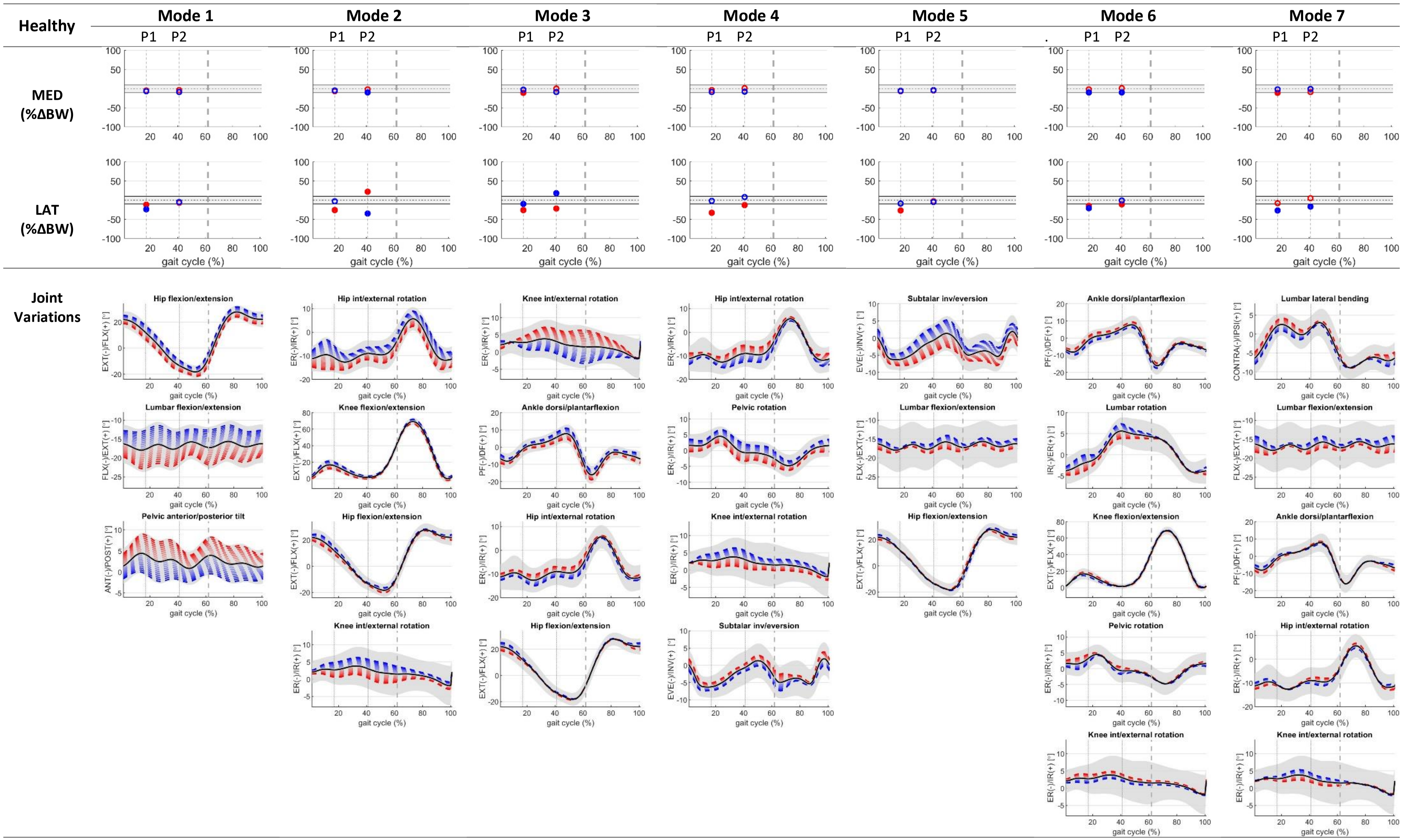
Healthy
Mode 1
Mode 2
Mode 3
Mode 4
Mode 5
Mode 6
Mode 7
P1
P2
MED (%ΔBW)
LAT (%ΔBW)
gait cycle (%)
Joint Variations
Hip flexion/extension
Lumbar flexion/extension
Pelvic anterior/posterior tilt
Hip int/external rotation
Knee flexion/extension
Knee int/external rotation
Ankle dorsi/plantarflexion
Pelvic rotation
Subtalar inv/eversion
Lumbar rotation
Lumbar lateral bending
EXT(-)/FLX(+) [°]
FLX(-)/EXT(+) [°]
ANT(-)/POST(+) [°]
ER(-)/IR(+) [°]
PF(-)/DF(+) [°]
EVE(-)/INV(+) [°]
IR(-)/ER(+) [°]
CONTRA(-)/IPSI(+) [°]

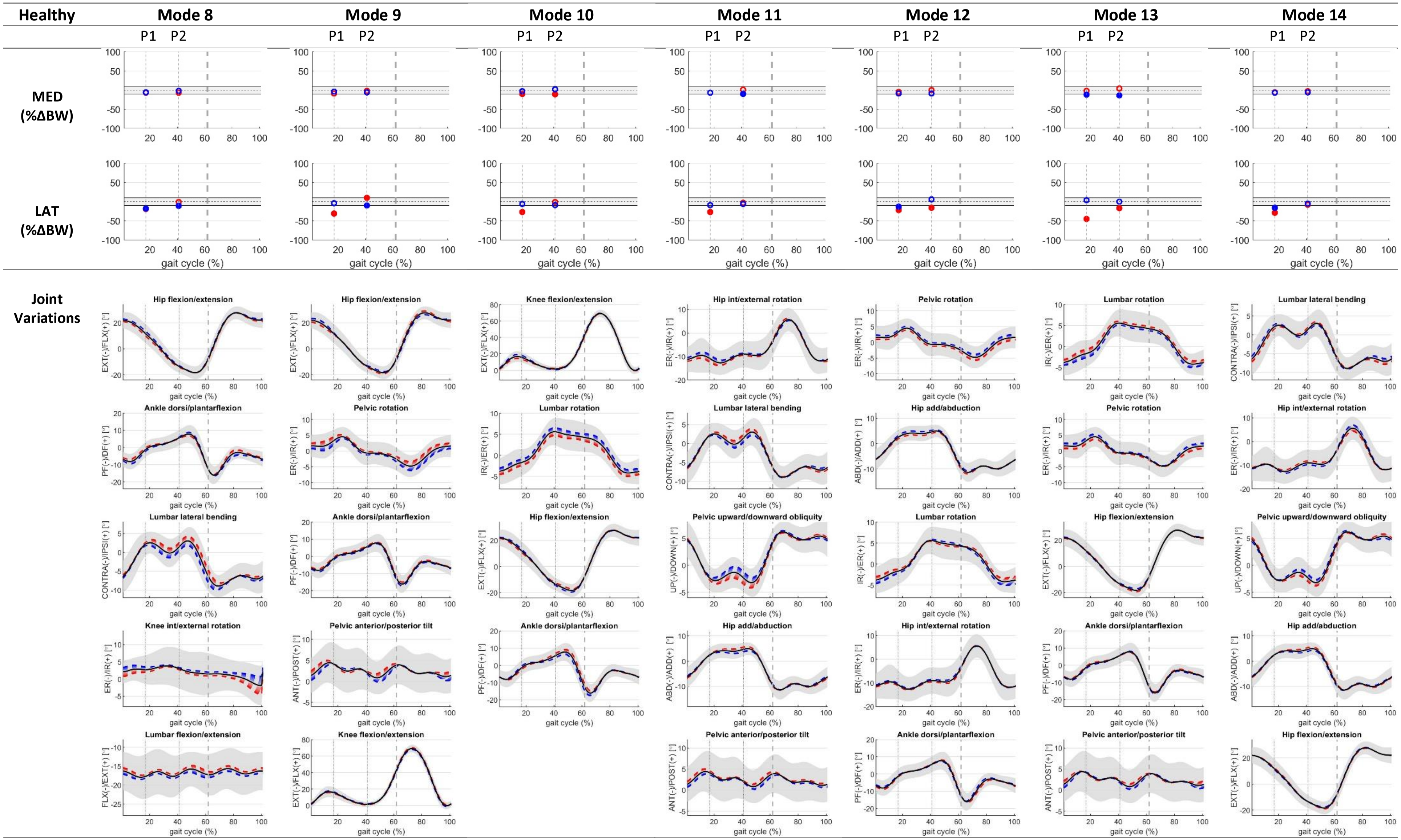

Healthy
Mode 8
Mode 9
Mode 10
Mode 11
Mode 12
Mode 13
Mode 14
P1
P2
MED (%ΔBW)
LAT (%ΔBW)
gait cycle (%)
Joint Variations
Hip flexion/extension
Knee flexion/extension
Hip int/external rotation
Pelvic rotation
Lumbar rotation
Lumbar lateral bending
Ankle dorsi/plantarflexion
Hip add/abduction
Pelvic upward/downward obliquity
Knee int/external rotation
Pelvic anterior/posterior tilt
Lumbar flexion/extension
EXT(-)/FLX(+) [°]
PF(-)/DF(+) [°]
CONTRA(-)/IPSI(+) [°]
ER(-)/IR(+) [°]
FLX(-)/EXT(+) [°]
ANT(-)/POST(+) [°]
IR(-)/ER(+) [°]
ABD(-)/ADD(+) [°]
UP(-)/DOWN(+) [°]

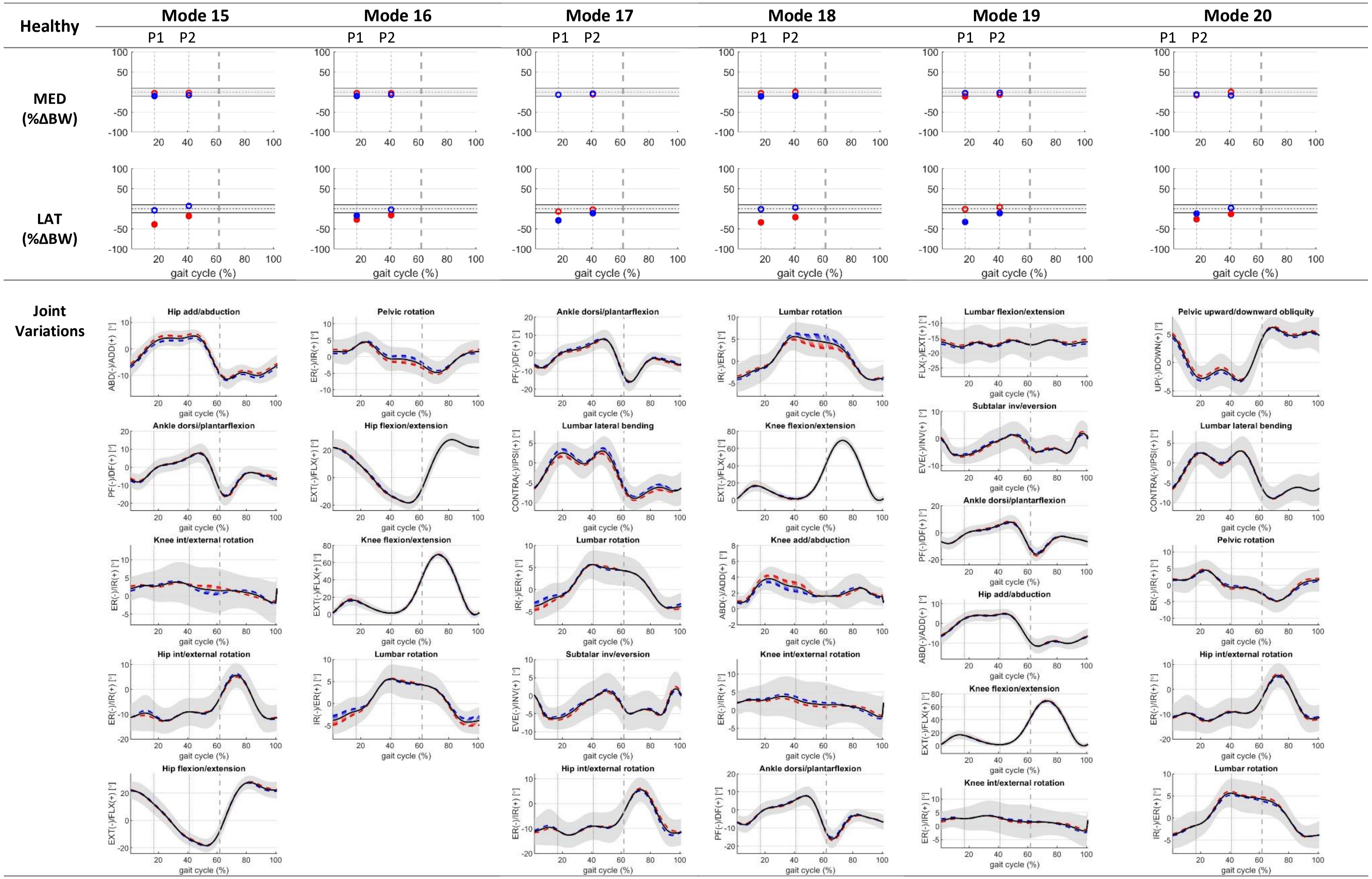

Healthy
Mode 15
Mode 16
Mode 17
Mode 18
Mode 19
Mode 20
P1
P2
MED (%ΔBW)
LAT (%ΔBW)
gait cycle (%)
Joint Variations
Hip add/abduction
Ankle dorsi/plantarflexion
Knee int/external rotation
Hip int/external rotation
Hip flexion/extension
Pelvic rotation
Knee flexion/extension
Lumbar rotation
Lumbar lateral bending
Subtalar inv/eversion
Knee add/abduction
Lumbar flexion/extension
Pelvic upward/downward obliquity
ABD(-)/ADD(+) [°]
PF(-)/DF(+) [°]
ER(-)/IR(+) [°]
EXT(-)/FLX(+) [°]
IR(-)/ER(+) [°]
CONTRA(-)/IPSI(+) [°]
EVE(-)/INV(+) [°]
FLX(-)/EXT(+) [°]
UP(-)/DOWN(+) [°]

***Figure S2***: *Changes as result of kinematic variations in medial and lateral compartment (relevant changes are depicted with filled red and blue circle) expressed as a % difference in body weight (BW) compared to knee contact forces estimated using the mean healthy gait pattern (black dotted lines), at the top. Solid grey lines are the cut-off/threshold for functionally relevant knee contact force changes (see Methods section). Below (bottom) are the kinematics of the different joints contributing to the top 50% (see Fig 4) of the observed variation for each specific mode depicted as ±1 STD (red, blue respectively). Grey dashed line defines the stance phase.*

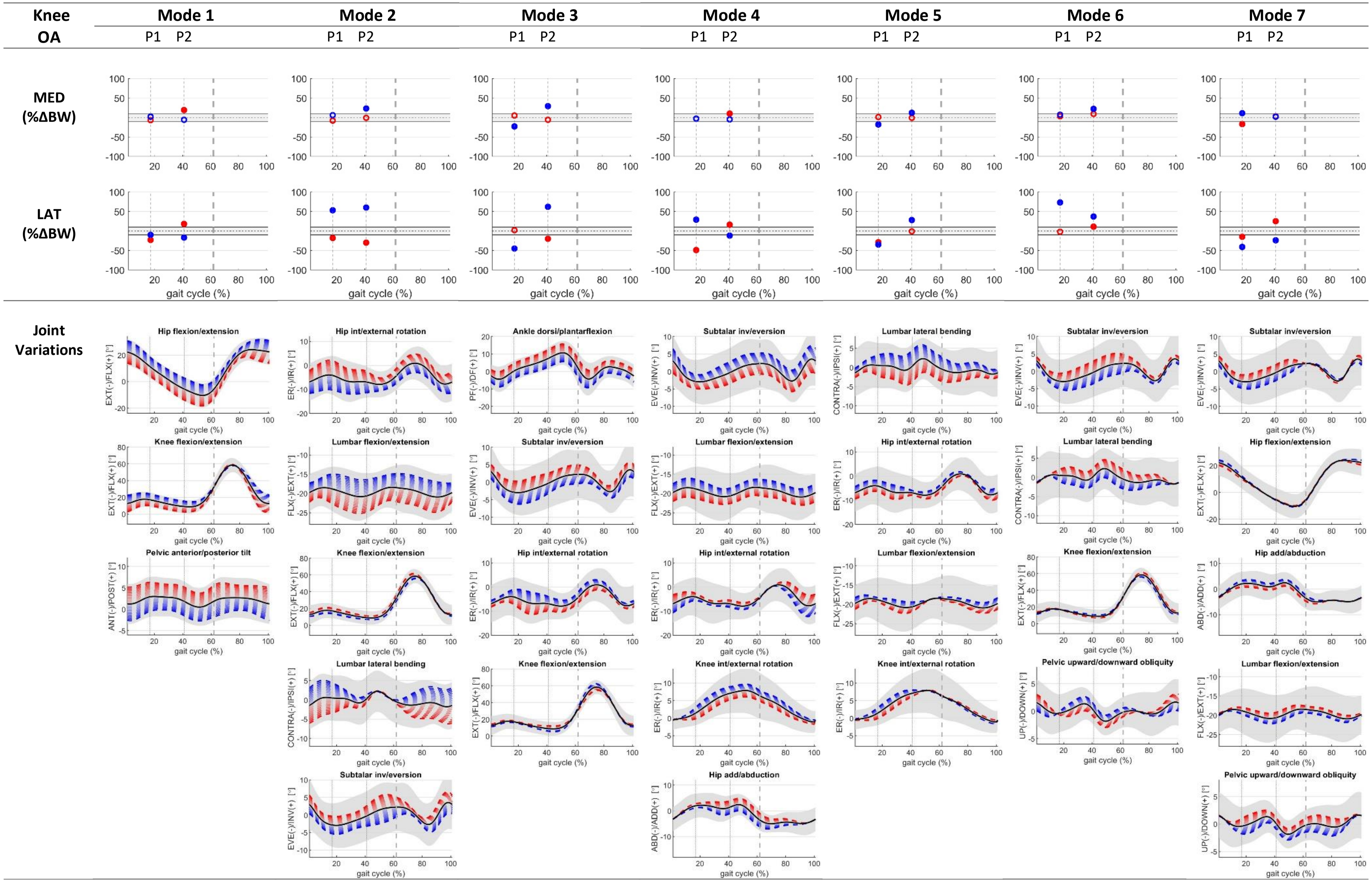

Knee OA
Mode 1
Mode 2
Mode 3
Mode 4
Mode 5
Mode 6
Mode 7
P1 P2
MED (%ΔBW)
LAT (%ΔBW)
gait cycle (%)
Joint Variations
Hip flexion/extension
Knee flexion/extension
Pelvic anterior/posterior tilt
Hip int/external rotation
Lumbar flexion/extension
Lumbar lateral bending
Subtalar inv/eversion
Ankle dorsi/plantarflexion
Knee int/external rotation
Hip add/abduction
Pelvic upward/downward obliquity
EXT(-)/FLX(+) [°]
ANT(-)/POST(+) [°]
ER(-)/IR(+) [°]
FLX(-)/EXT(+) [°]
CONTRA(-)/IPSI(+) [°]
EVE(-)/INV(+) [°]
PF(-)/DF(+) [°]
ABD(-)/ADD(+) [°]
UP(-)/DOWN(+) [°]

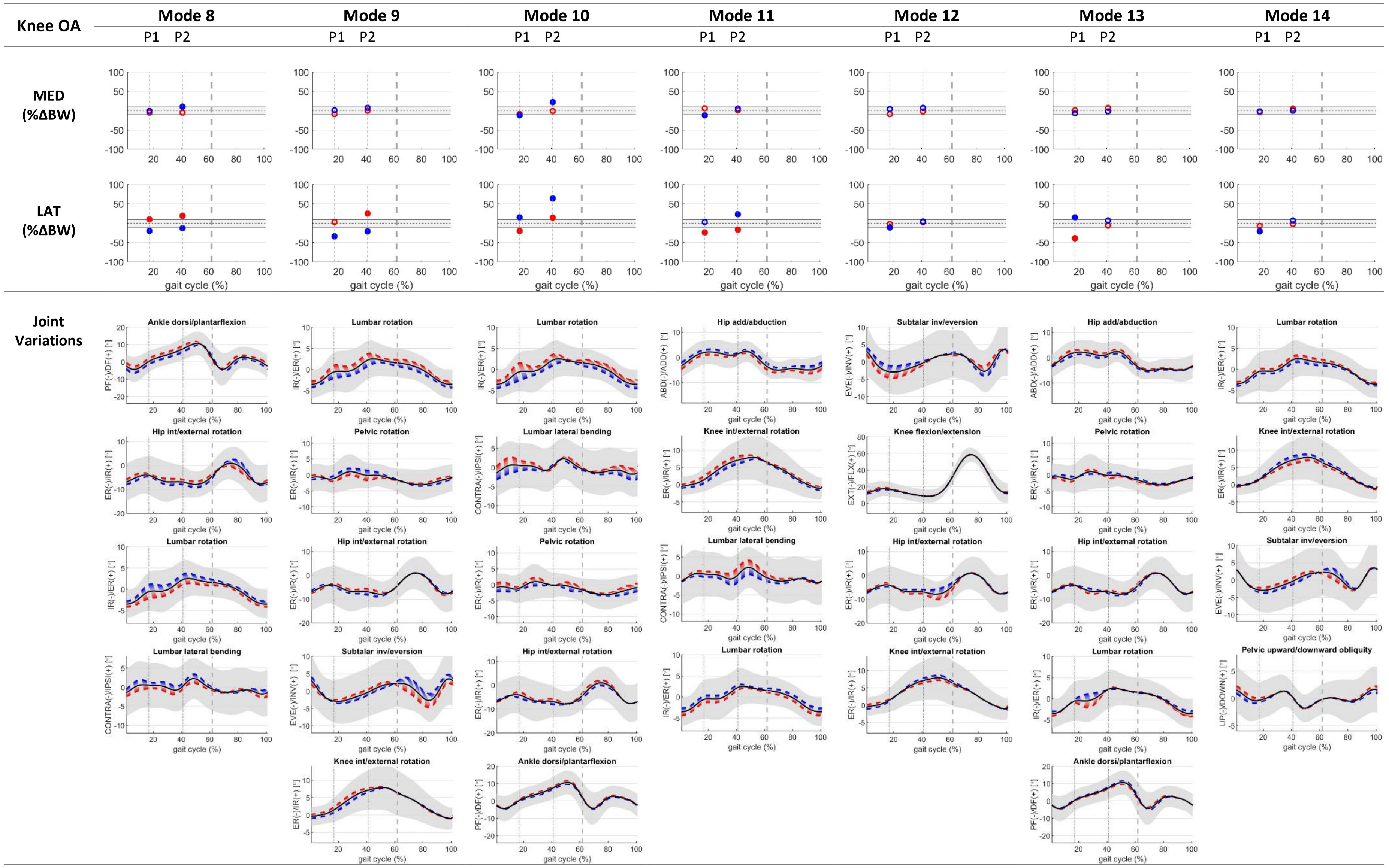

Knee OA
Mode 8
Mode 9
Mode 10
Mode 11
Mode 12
Mode 13
Mode 14
P1 P2
MED (%ΔBW)
LAT (%ΔBW)
Joint Variations
gait cycle (%)
Ankle dorsi/plantarflexion
Hip int/external rotation
Lumbar rotation
Lumbar lateral bending
Pelvic rotation
Subtalar inv/eversion
Knee int/external rotation
Hip add/abduction
Knee flexion/extension
Pelvic upward/downward obliquity
PF(-)/DF(+) [°]
ER(-)/IR(+) [°]
IR(-)/ER(+) [°]
CONTRA(-)/IPSI(+) [°]
EVE(-)/INV(+) [°]
ABD(-)/ADD(+) [°]
EXT(-)/FLX(+) [°]
UP(-)/DOWN(+) [°]

*Figure S3: Changes as result of kinematic variations in medial and lateral compartment (relevant changes are depicted with filled red and blue circle) expressed as a % difference in body weight (BW) compared to knee contact forces estimated using the mean knee OA gait pattern (black dotted lines), at the top. Solid grey lines are the cut-off/threshold for functionally relevant knee contact force changes (see Methods section). Below (bottom) are the kinematics of the different joints contributing to the top 50% (see Fig 4) of the observed variation for each specific mode depicted as ±1 STD (red, blue respectively). Grey dashed line defines the stance phase.*

**Healthy:**

**Gait primitives/kinematics affecting lateral compartment loading:**

No relevant **increases** (>10%BW) in the 1st peak were observed in any of the modes. Increases in the 2nd peak were observed in mode 2 and 3, representing variation in (2) hip external rotation, knee extension, hip flexion and knee external rotation; (3) knee external rotation, ankle dorsiflexion, hip external rotation and hip extension.

Relevant **decreases** (<-10%BW) for either 1$^{st}$ and 2$^{nd}$ peaks were observed in mode 3, 4, 6, 12, 13, 15, 16, 18, and 2. These modes represent variations in (3) knee internal rotation, ankle plantarflexion, internal rotation and hip extension; (4) hip internal rotation, pelvic internal rotation, knee external rotation and subtalar inversion; (6) ankle dorsiflexion, lumbar internal rotation, knee extension, pelvic external rotation and knee internal rotation; (12) pelvic internal rotation, hip abduction, lumbar internal rotation, hip external rotation and ankle plantarflexion; (13) lumbar external rotation, pelvic internal rotation, hip flexion, ankle plantarflexion and pelvic anterior tilt; (15) hip adduction, ankle dorsiflexion, knee external rotation, hip external rotation and hip extension; (16) pelvic internal rotation, hip flexion, and lumbar internal rotation; (18) lumbar external rotation, knee extension, knee adduction, knee external rotation and ankle plantarflexion; (20) pelvic upward obliquity, lumbar contralateral bending, pelvic external rotation, hip external rotation and lumbar external rotation. Decreases in the 1st peak only were observed in mode 1, 5, 8, 9, 10, 11 and 14. These modes represent variations in (1) hip flexion, lumbar extension and pelvic anterior tilt; (5) subtalar eversion, lumbar flexion and hip extension; (8) hip flexion, ankle plantarflexion, lumbar contralateral bending, knee internal rotation and lumbar flexion; (9) hip extension, pelvic external rotation, ankle dorsiflexion, pelvic posterior tilt and knee flexion; (10) knee extension, lumbar internal rotation, hip flexion and ankle plantarflexion; (11) hip external rotation, lumbar contralateral bending, pelvic downward obliquity, hip abduction and pelvic anterior tilt; (14) lumbar contralateral bending, hip internal rotation, pelvic upward obliquity, hip abduction and hip flexion. Decreases in the 2nd peak only were observed in mode 2: hip internal rotation, knee flexion, hip flexion and knee internal rotation.

**Knee OA:**

**Gait primitives/kinematics affecting lateral compartment loading:**

Relevant **increases** for either 1$^{st}$ and 2$^{nd}$ peaks were observed in mode 2, 6, 10. These modes represent variations in (2) hip external rotation, lumbar extension, knee extension, lumbar ipsilateral bending and subtalar eversion; (6) subtalar eversion, lumbar contralateral bending, knee flexion and pelvic upward obliquity; (10) lumbar internal rotation, lumbar ipsilateral bending, pelvic external rotation, hip external rotation and ankle plantarflexion. Increases in the 1$^{st}$ peak only were observed in mode 4 and 13: (4) subtalar inversion, lumbar extension, hip internal rotation, knee internal rotation and hip abduction; (13) hip abduction, pelvic external rotation, lumbar external rotation and ankle dorsiflexion. And increases in the 2$^{nd}$ peak only were observed in mode 1, 3, 4, 5, 7, 9 and 11. These modes represent variations in (1) hip extension, knee extension and pelvic posterior tilt; (3) ankle plantarflexion, subtalar eversion, hip internal rotation and knee extension; (4) subtalar eversion, lumbar flexion, hip external rotation, knee external rotation and hip adduction; (5) lumbar ipsilateral bending, hip internal rotation, lumbar extension and knee internal rotation; (7) subtalar inversion, hip extension, hip abduction, lumbar extension and pelvic upward obliquity; (9) lumbar external rotation, pelvic internal rotation, hip internal rotation, subtalar inversion and knee internal rotation; (11) hip adduction, knee external rotation, lumbar contralateral bending and lumbar external rotation.

Relevant **decreases** for either 1$^{st}$ and 2$^{nd}$ peaks were observed in mode 2, 7, 8, 9, 11. These modes represent variations in (2) hip internal rotation, lumbar flexion, knee flexion, lumbar contralateral bending and subtalar inversion; (7) subtalar eversion, hip extension, hip adduction, lumbar flexion and pelvic downward obliquity; (9) lumbar internal rotation, pelvic external rotation, hip external rotation, subtalar eversion and knee external rotation; (11) hip abduction, knee internal rotation, lumbar ipsilateral bending and lumbar internal rotation. Decreases in the 1st peak only were observed in mode 1, 3, 4, 5, 7, 10, 12, 13, 14. These modes represent variations in (1) hip extension, knee extension and pelvic posterior tilt; (3) ankle plantarflexion, subtalar eversion, hip internal rotation and knee extension; (4) subtalar eversion, lumbar flexion, hip external rotation, knee external rotation and hip adduction; (5) lumbar ipsilateral/contralateral bending, hip internal/external rotation, lumbar extension/flexion and knee internal/external rotation; (7) subtalar inversion, hip extension, hip abduction, lumbar extension and pelvic upward obliquity; (10) lumbar external rotation, lumbar ipsilateral bending, pelvic external rotation, hip internal rotation and ankle dorsiflexion; (12) subtalar inversion, knee extension, hip external rotation and knee external rotation; (14) lumbar internal rotation, knee internal rotation, subtalar eversion and pelvic downward obliquity. Decreases in the 2st peak only were observed in mode 3 and 4. These modes represent variations in (3) ankle dorsiflexion, subtalar inversion, hip external rotation and knee flexion; (4) subtalar inversion, lumbar extension, hip internal rotation, knee internal rotation and hip abduction.

**Healthy**

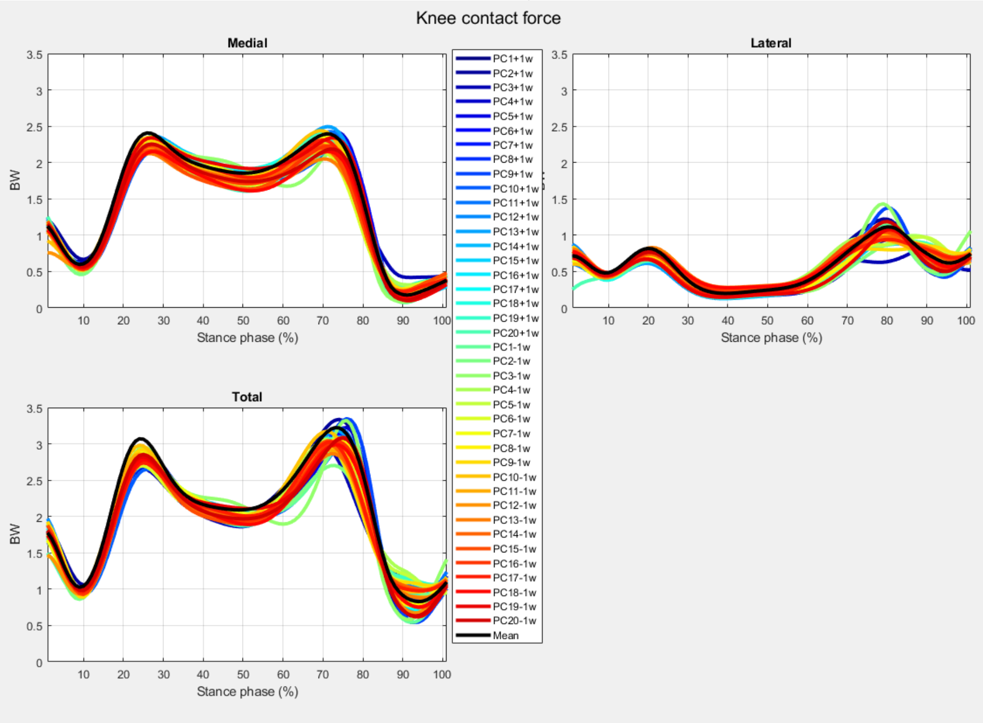


**Knee OA**

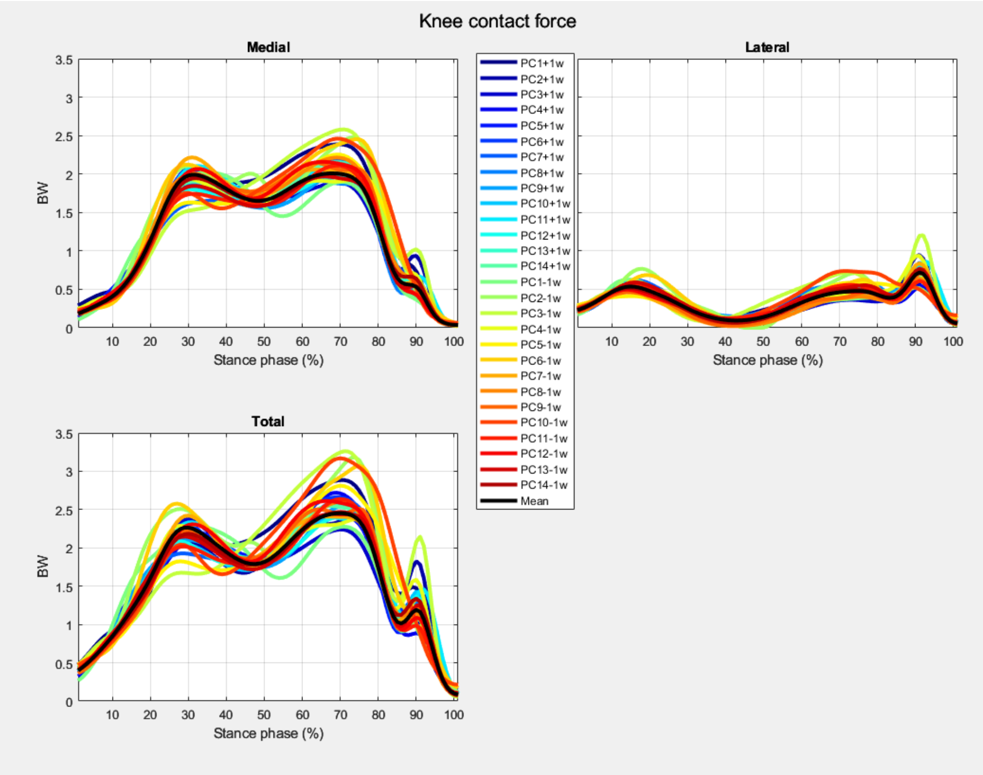


***Figure S4**: Knee contact force variations (total, medial, and lateral compartments) for the healthy population (top figure) and knee OA population (bottom figure) are represented by different modes (PC) - colored solid lines - in comparison to the mean knee contact forces - black solid line.*
